# Integrative analysis of GWAS and co-localisation data suggests novel genes associated with age-related multimorbidity

**DOI:** 10.1101/2022.11.11.22282236

**Authors:** Clare E. West, Mohd Karim, Maria J. Falaguera, Leo Speidel, Charlotte J. Green, Lisa Logie, Jeremy Schwartzentruber, David Ochoa, Janet M. Lord, Michael A. J. Ferguson, Chas Bountra, Graeme F. Wilkinson, Beverley Vaughan, Andrew R. Leach, Ian Dunham, Brian D. Marsden

## Abstract

Advancing age is the greatest risk factor for developing multiple age-related diseases. When developing therapeutics, using a Geroscience approach to target the shared underlying pathways of ageing, rather than individual diseases, may be an effective way to treat and prevent age-related morbidity while potentially reducing the burden of polypharmacy. We harness the Open Targets Platform and Open Targets Genetics Portal to perform a systematic analysis of nearly 1,400 genome-wide association studies (GWAS) mapped to 34 age-related diseases and traits to identify genetic signals that appear to be shared between two or more of these traits. We identify 995 targets with shared genetic links to these age-related diseases and traits, which are enriched in mechanisms of ageing and include known ageing and longevity-related genes. Of these 995 genes, 128 are the target of an approved or investigational drug, 526 have experimental evidence of binding pockets or are predicted to be tractable by small molecule or antibody modality approaches, and 341 have no existing tractability evidence, representing underexplored genes which may reveal novel biological insights and therapeutic opportunities. We present these candidate targets in a web application, TargetAge, to enable the exploration and prioritisation of possible novel drug targets for age-related multimorbidity.

## Introduction

Life expectancy is increasing globally, bringing with it the need for new approaches and therapeutics to promote healthy ageing (Partridge, Deelen, and Slagboom 2018). For instance, the UK Office for National Statistics reported that the proportion of the population aged over 85 is projected to increase from 2.5% in 2020 to 4.3% by 2050 (Office for National Statistics 2022). However, the number of years in good health -- the “healthspan” – is not increasing at the same pace (Great Britain. Parliament. House of Lords. Science and Technology Committee 2021; Partridge, Deelen, and Slagboom 2018) and people are experiencing an increasing number of years of ill-health in later life. More than half of people over 65 in the UK suffer from multiple long-term health conditions (NIHR Dissemination Centre 2018) known as multimorbidity, for which it is common for people to be taking five or more medications, known as polypharmacy (Masnoon et al. 2017). The traditional model of tackling diseases independently during research, drug development, clinical trials, and treatment may not be best serving these patients, who represent an increasingly large proportion of the general population. Advancing age is the greatest risk factor for developing multimorbidity, suggesting that biological ageing processes play a pathogenic role. A Geroscience approach, one that aims at tackling combinations of diseases via their common underlying ageing pathways, has been proposed as a way to treat and prevent age-related morbidity more effectively whilst potentially reducing the burden of polypharmacy (Chaib, Tchkonia, and Kirkland 2022; Ermogenous et al. 2020; Whitty et al. 2020).

Ageing is associated with time-dependent functional decline, initially manifesting as subclinical physiological changes in the body, progressing to more systemic changes, and ultimately defined by disease conditions and multimorbidity (Partridge, Deelen, and Slagboom 2018). The rate of decline relative to chronological age varies between individuals, and there is evidence from animal models and humans that this can be modified through interventions (Ermogenous et al. 2020; Partridge, Deelen, and Slagboom 2018). Specific biological processes are thought to contribute to ageing and the pathology of age-related diseases, termed the Hallmarks of Ageing (Lopez-Otin et al. 2013), and comprise: Telomere shortening, epigenetic modifications, genomic instability, reduced proteostasis, altered nutrient sensing, mitochondrial dysfunction, cell senescence, stem cell exhaustion and altered intercellular communication, with increased systemic inflammation a key component of the latter.

In support of generic mechanisms driving multimorbidity, previous genetic studies have found that age-related diseases are genetically correlated, and that genes with multi-trait associations often relate to biological processes that are implicated in ageing. Belloy *et al* (Belloy, Napolioni, and Greicius 2019) found significant genetic overlap between 8 age-related diseases and parental lifespan, based on 961 genome-wide significant variant-trait associations in the GWAS catalog. The authors identified 12 multi-trait loci, including variants relating to inflammation, obesity, blood lipid levels, DNA repair mechanisms, and telomere maintenance. For instance, Apolipoprotein E (APOE) is involved in lipid transport and neural health, and is strongly linked to multiple age-related traits including Alzheimer’s disease, cardiovascular disease, stroke, and longevity (Belloy, Napolioni, and Greicius 2019).

Dönertaş *et al* (Dönertaş et al. 2021) identified common genetic associations between age-related diseases using a data-driven approach. Using self-reported disease data for UK Biobank (UKBB) participants aged up to 70 years, diseases were clustered based on age-of-onset profiles. One cluster of diseases showed a rapid increase in incidence after age 40, and contained cardiovascular diseases, diabetes, osteoporosis and cataracts. A second cluster shared a slower age-related rate of increase from age 20, including musculoskeletal and gastrointestinal diseases, anaemia, deep vein thrombosis, thyroid problems and depression. Two final clusters contained diseases with childhood-onset (primarily inflammatory diseases) and uniformly distributed age-of-onset (respiratory and infectious diseases). GWAS performed on each disease found a high level of genetic similarity within clusters of diseases with similar age-of-onset profiles, even after controlling for shared disease categories and co-occurrences. Furthermore, the diseases in clusters with age-dependent onset profiles were associated with 581 genes, which were significantly enriched in known ageing-, longevity-, and senescence-related genes.

Pun *et al* (Pun et al. 2022) used deep learning models including omics, literature, and key opinion leader scores from multiple data sources to predict a list of 484 genes implicated in a set of 14 age-associated diseases. Well-known age-related genes were found, along with a number of novel genes. A final set of 9 genes was prioritised with strong links to inflammation and epigenetic programming.

Whilst this biological relationship between ageing and multimorbidity suggests a potential strategy for therapeutic intervention via shared pathways, translating this biological understanding to clinical benefits in patients presents a challenge (Chaib, Tchkonia, and Kirkland 2022). Although a number of promising novel drug targets have been explored for the treatment of multimorbidity, as well as repurposing existing drugs such as Metformin (Barzilai et al. 2016; Dönertaş et al. 2018), there has so far been limited success in clinical trials (Ermogenous et al. 2020).

Here we harness publicly available resources for systematic drug target identification and prioritisation for individual diseases, as well as high-quality manually curated databases of genes and drugs related to ageing and longevity, to produce a set of potentially tractable targets for ageing-related comorbidities and multimorbidity. As human genetic evidence has been shown to improve the odds of successful drug discovery (King, Davis, and Degner 2019), we focus on a genetics approach to identifying potential drug targets implicated in multiple ageing-related diseases. We use the Open Targets Platform (Ochoa et al. 2020) and Open Targets Genetics Portal (Ghoussaini et al. 2021) to perform a systematic analysis of 1,394 genome-wide association studies (GWAS) mapped to a curated list of age-related diseases and traits to identify genetic signals that appear to be shared between two or more of these traits. We map these signals to the most likely candidate causal genes and assess these genes in terms of existing links to ageing and actionability as possible drug targets. These include genes with demonstrated longevity-altering effects in model organisms or links to ageing-related pathways, as well as a large number of novel genes, many of which are predicted to be tractable drug targets. Finally, we integrate these data in TargetAge, a web application to enable the identification and prioritisation of possible novel drug targets for age-related multimorbidity.

## Results

### GWAS studies for age-related diseases and biomarkers

An overview of the genetic analysis is illustrated in Fig 1. We curated a list of 34 chronic age-related diseases and biomarkers that correlate with ageing and multimorbidity (such as grip strength measurement and gait speed) (SI Table 1). We defined these using experimental factor ontology (EFO) terms (Malone et al. 2010), and included relevant descendant terms for these diseases (e.g. in addition to “osteoarthritis” we also included the child term “osteoarthritis, hip”). Terms categorised as genetic, familial or congenital disease (i.e. inherited from parents typically manifesting at, before, or just after birth), as well as pregnancy or perinatal diseases were removed as these are less likely to be ageing-related traits. We additionally excluded cell proliferation disorders; whilst some cancers are prevalent amongst older people, cancers have distinct genetic and environmental disease pathways that are not in the scope of the present study. A full list of traits, corresponding EFO codes, and descendant terms are included in SI Table 2.

**Figure 1.**
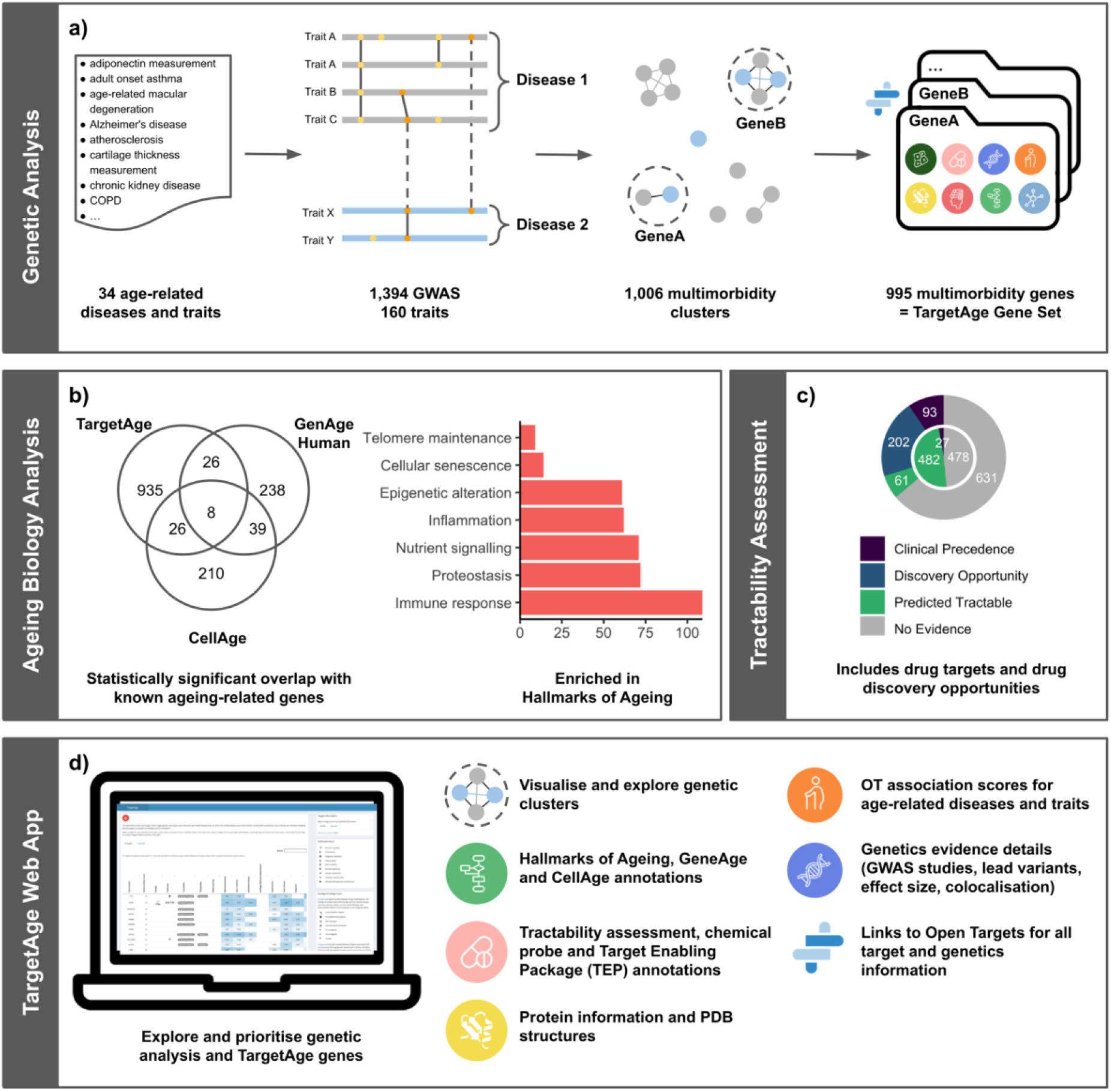
Overview of the TargetAge analysis. a) **Genetic Analysis:** GWAS data from Open Targets Genetics is retrieved for 34 age-related diseases and traits and descendant terms in the Experimental Factor Ontology (EFO). Overlapping genetic hits that may share a causal variant are clustered to identify independent genetic signals. Clusters containing multiple age-related diseases are mapped to candidate causal genes using the L2G method from Open Targets Genetics. Target annotations are retrieved from Open Targets Platform for the 995 resulting protein coding genes, referred to as the TargetAge gene set. b) **Ageing Biology Analysis**: The TargetAge set has statistically significant overlaps with three sets of age-related genes: GenAge, CellAge, and genes annotated with GO terms mapped to the Hallmarks of Ageing. c) **Tractability Assessment**: The TargetAge set were assessed for clinical precedence and tractability as drug targets using a therapeutic antibody (outer ring) or small molecule (inner) approach, as well as for the availability of chemical probes and Target Enabling Packages (TEPs). d) **TargetAge Web App**: The TargetAge gene set are presented in a freely available web application including the genetic evidence, target annotations, links to ageing, and tractability assessment.

We retrieved all GWAS studies included in Open Targets Genetics (https://genetics.opentargets.org/) with a sample size (or number of cases, for case-control studies) of at least 2,000 for which the reported traits mapped to these age-related traits of interest, or their descendants in the ontology. This resulted in 1,394 GWAS studies representing 160 different traits mapping to 30 of the 34 curated age-related traits (SI Table 1). The total number of studies available for each trait ranged from 1 (idiopathic pulmonary fibrosis, chronic pain, and cartilage thickness measurement) to 649 (lipid measurement) with a median of 18.5. Most studies were carried out in populations with European (n=828, 59.4%), majority European (n=268, 19.2%) or East Asian (n=107, 7.7%) ancestry. For these studies, we retrieved the lead variants from each locus with a genome-wide significant association (P < 5 × 10^−8^).

### Clustering overlapping GWAS hits identifies independent genetic associations

To estimate the number of independent genetic signals for the ageing-related traits, we clustered together GWAS hits that may share a causal variant. To define potentially shared signals, we used co-localisation evidence where summary statistics were available (requiring a posterior probability of co-localisation greater than 0.8), otherwise overlap of variants derived from Linkage Disequilibrium (LD) expansion or fine mapping (see Methods).

Around 12% of hits (n=3,524) were only identified in a single GWAS. The remaining associations were grouped into 3,158 clusters of which 2,152 contained associations to one trait and 1,006 clusters involved more than one trait.

This approach aims to cluster together GWAS hits that correspond to a single genetic signal. However, in some cases, the resulting clusters containing multiple associations may still include multiple distinct genetic signals. To investigate whether clusters could be subdivided further, we applied, to each cluster, a community detection algorithm that aims to identify subclusters with weak co-localisation or LD evidence across subclusters and strong evidence within subclusters (see Methods). Most clusters consisted of just one community (77% of all clusters, 49% of multi-trait clusters) or two communities (19% of all clusters, 39% of multi-trait clusters). The largest number of communities within a single cluster was 9 (cluster ID 21) consisting of 299 nodes from 15 traits and was most strongly associated with the gene encoding APOE.

### Shared clusters identify genetic correlations between related traits

Using these clusters of genetic associations to age-related traits, we tested whether the overlap in genetic signals between pairs of traits was larger than expected by chance given the total number of genetic signals linked to each trait (one-sided Fisher’s exact test, see Methods) (Fig 2). Overall, 96 of the 406 combinations have a p-value < 0.05, of which 55 combinations are significant after adjusting for multiple testing (see Methods). This suggests that the set of ageing-related diseases and traits are enriched with genetically correlated traits.

**Figure 2.**
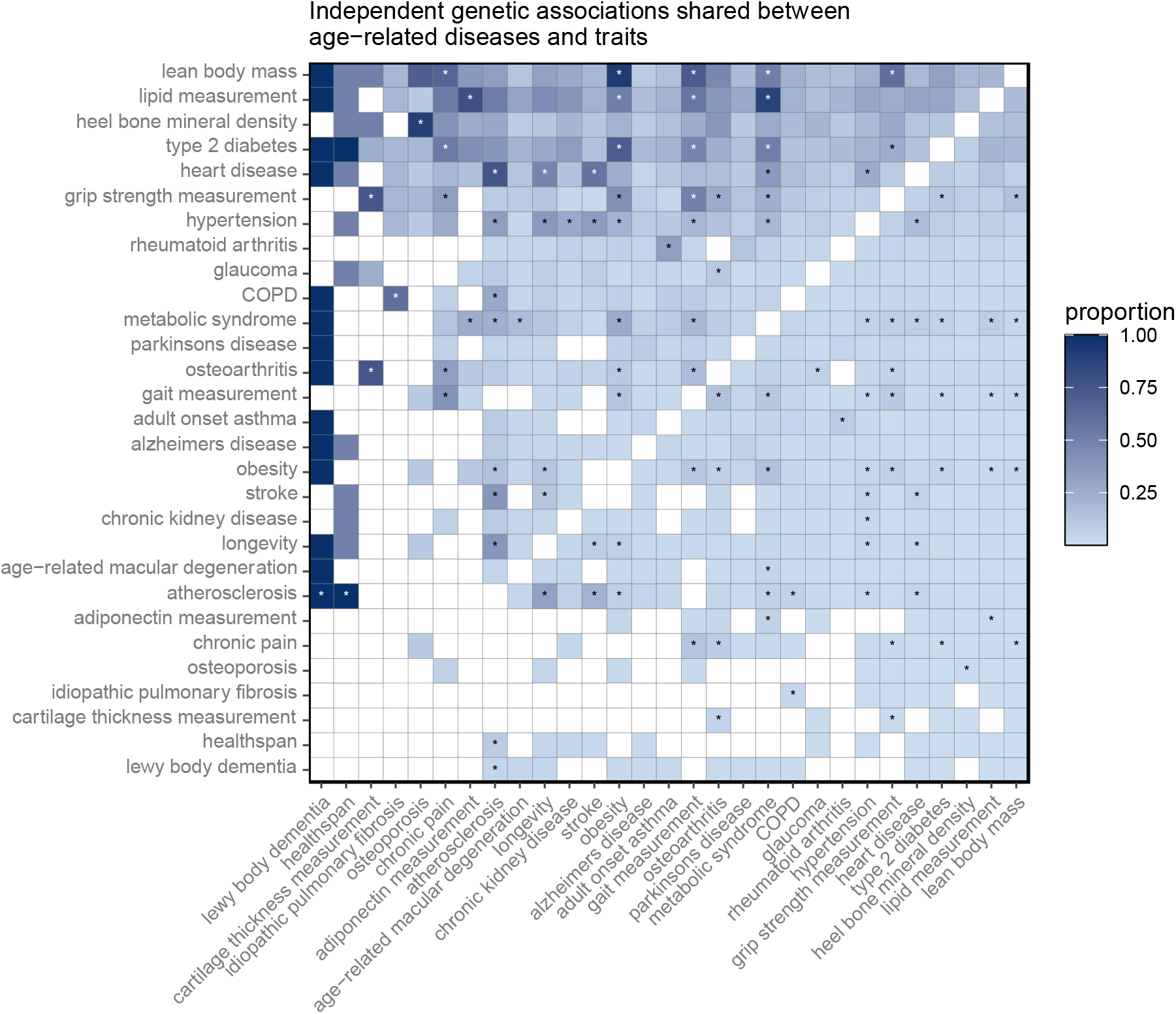
The proportion of independent genetic associations shared between age-related diseases and traits. Proportions are relative to the total number of independent genetic associations identified for the trait on the horizontal axis; darker blue indicates a greater proportion of shared genetic associations. Cells marked with an asterisk are statistically significant after adjusting for multiple testing of all possible pairwise combinations of traits. Traits are ordered by the total number of implicated genetic association clusters. See SI Fig 2 for the number of overlaps between each combination.

Most of the significant overlaps are consistent with previously identified relationships between ageing-related diseases and traits, indicating that the clustering method captures relationships between correlated traits. The most significant overlap is between lipid measurement and metabolic syndrome. Metabolic syndrome is a combination of metabolic abnormalities including altered blood lipid levels, hypertension, obesity, and insulin resistance, which are risk factors for type 2 diabetes and cardiovascular disease (Rochlani et al. 2017). As expected, metabolic syndrome has significant overlaps with all these related traits, as well as age-related macular degeneration, gait measurement, and grip strength measurement. The next most significant overlap is between lean body mass and grip strength measurement, the diagnostic measurements used to define sarcopenia, a common ageing-related condition characterised by loss of skeletal muscle mass, strength and function (Cruz-Jentoft et al. 2019). This is followed by the highly correlated traits lean body mass and obesity, as well as type 2 diabetes and obesity. Other overlapping traits that reflect known clinical associations include hypertension with chronic kidney disease and type 2 diabetes (Sun et al. 2019; Tatsumi and Ohkubo 2017), and age-related macular degeneration with metabolic syndrome and type 2 diabetes. There is also a significant overlap between loci associated with heel bone mineral density and osteoporosis. Gait measurement and grip strength measurement significantly overlap and are known to negatively correlate with multimorbidity (Calderón-Larrañaga et al. 2019; Bohannon 2019).

We find a significant overlap between adult-onset asthma and rheumatoid arthritis; some epidemiological studies have found a positive association between asthma and rheumatoid arthritis, but the relationship is poorly understood (Rolfes et al. 2017; S. Y. Kim et al. 2019). We also find a significant overlap between osteoarthritis and glaucoma. While inflammatory forms of arthritis are thought to increase the risk of glaucoma due to intraocular pressure, and a possible common autoimmune-mediated pathological pathway has been proposed for rheumatoid arthritis and glaucoma (S. H. Kim et al. 2022; Geyer and Levo 2020), there is no established link between osteoarthritis and glaucoma.

The single lead variant associated with Lewy body dementia (19_44908684_T_C, rs429358) is part of cluster 21 that is linked to *APOE* and involves 9 age-related traits including age-related macular degeneration, Alzheimer’s disease, atherosclerosis, as well as longevity. Longevity has significant overlaps with healthspan, atherosclerosis, stroke, obesity, osteoarthritis, type 2 diabetes, and lipid measurement.

These results are consistent with previous findings on the shared underlying genetics of ageing-related diseases and traits. We hypothesise that genes implicated with multiple ageing-related traits could represent shared aetiology that may relate to underlying ageing processes.

### Shared GWAS associations between traits implicate 995 genes in multiple age-related morbidities

We mapped each independent genetic association cluster to possible causal genes using the Open Targets Genetics ‘locus to gene’ (L2G) score for each locus in the cluster (Mountjoy et al. 2021) (Methods). Of the 1,006 clusters involving more than one trait, 796 (79%) contain at least one predicted causal gene (L2G score >= 0.5) with 178 of these supported by co-localisation with protein quantitative trait loci (pQTL) or gene expression quantitative trait loci (eQTL). Overall, there are 1,021 predicted causal genes linked to at least one multimorbidity cluster, of which 995 are protein-coding, which we refer to as the “TargetAge” gene set. Overall, 290 (29%) of TargetAge genes are annotated to at least one Hallmark of Ageing (*p* = 1.40 × 10^−6^).

The majority of TargetAge genes are linked to one (n=905, 91%) or two clusters (n=80, 8%). There are eight genes (*MPPED2, APOB, LDLR, TBX3, ZFHX3, BCL2L11, SMAD3*, and *IRS1*) linked to three clusters, and two genes, *PPARG* and *LRMDA*, are linked to four clusters. Of the genes linked to more than two clusters, *IRS1* (insulin receptor substrate 1) and *PPARG* (peroxisome proliferator activated receptor gamma) have previously been linked to ageing and are included in GenAge, a manually curated database of ageing-related genes due to longevity-altering effects in model organisms (Tacutu et al. 2018). Knockout of the gene homologous to *IRS1* resulted in increased lifespan in both mice (Selman et al. 2008) and drosophila (Clancy et al. 2001). *PPARG* is an important regulator of several ageing-related pathways and has genetic links to type 2 diabetes, atherosclerosis, and longevity; mice with lowered expression of the homologous gene have reduced lifespan (Argmann et al. 2009). A further four genes have links to the Hallmarks of Ageing, including *TBX3* (T-box transcription factor 3) a regulator of cellular senescence (Kumar P et al. 2014). *SMAD3* (SMAD family member 3) is a regulator of inflammation and an inhibitor of *PPARG* expression (Tan et al. 2004); decreased expression of the orthologue Smad3 has been linked to ageing, neuroinflammation, and neurodegeneration in mouse models (Liu et al. 2020). *LDLR* (low density lipoprotein receptor) and *APOB* (apolipoprotein B) are involved in proteostasis and nutrient signalling. *APOB* has been linked to longevity in humans and mice, and may play a role in the pro-longevity effects of dietary restriction (Mutlu, Duffy, and Wang 2021).

The remaining four genes that were linked to more than two clusters, but do not have established links to GenAge or the Hallmarks of Ageing are *MPPED2* (metallophosphoesterase domain containing 2), *ZFHX3* (zinc finger homeobox 3), *BCL2L11* (BCL2 like 11), and *LRMDA* (leucine rich melanocyte differentiation associated). Although it does not have a known role in inflammation, a *MPPED2* polymorphism has been associated with altered systemic inflammation and adverse outcomes trauma patients (Schimunek et al. 2019). The transcription factor *ZFHX3* is associated with atrial fibrillation, an ageing-related condition, and has a potential role in inflammation (Zaw et al. 2017; Tomomori et al. 2018). *LRMDA* is involved in melanocyte differentiation, and *BCL2L11* is a mediator of apoptosis and is associated with tumorigenesis (Dai et al. 2022).

### TargetAge genes overlap with known age-related genes and the Hallmarks of Ageing

We compared the TargetAge gene set with three sets of genes with known links to ageing. Two are manually curated databases of human genes linked to ageing or longevity (GenAge Human, 307 genes (Tacutu et al. 2018)), and cellular senescence (CellAge, 279 genes (Avelar et al. 2020)). The third is a set of 4,527 genes annotated with Gene Ontology (GO) biological process terms representing each of the Hallmarks of Ageing (see Methods).

The most highly represented hallmark is the immune inflammatory response (11% of genes, Figure 3a). The TargetAge set of multimorbidity genes also overlaps with both the GenAge Human and CellAge gene sets (Fig 1b). Of the 995 TargetAge genes, 34 (3.4%) are in GenAge Human (representing 11% of the GenAge Human database) (*p* = 1.4 × 10^−5^), and 34 (3.4%) are in CellAge (representing 12% of the CellAge database) (Fig 2b) (*p* = 2.0 × 10^−6^). Whilst these significant overlaps indicate shared biology coverage between TargetAge, GenAge, and CellAge, the sets are mostly distinct, which suggests that there are elements of unique biology that are being captured by each set.

**Fig 3.**
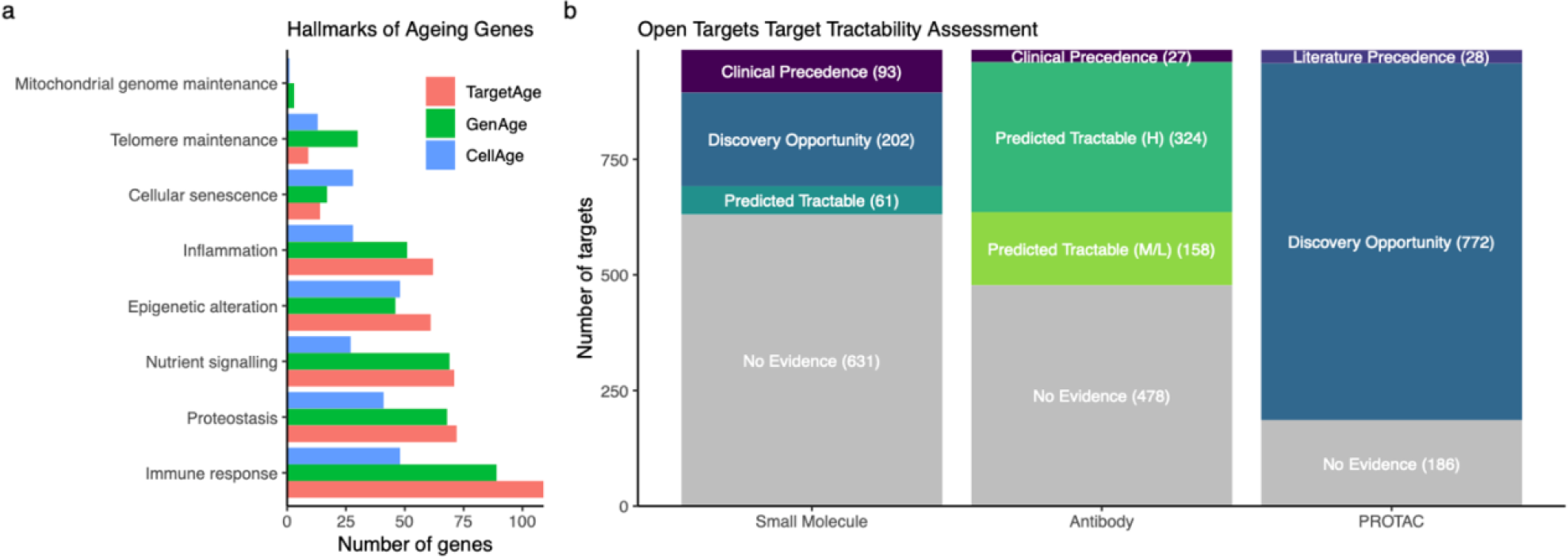
**a**) The proportion of genes annotated with at least one GO term corresponding to each Hallmarks of Ageing category, for the TargetAge gene set (red), GenAge (green) and CellAge (blue). **b)** Tractability categorisations when considering Small Molecule, Antibody, or PROTAC therapeutic approaches. Categorisations are based on the existence of an approved or experimental drug (Clinical Precedence), literature reports for PROTAC (Literature Precedence), and different levels of experimental and computational evidence.

### TargetAge genes include drug targets and drug development opportunities

TargetAge genes are implicated in multiple age-related diseases and are enriched in ageing processes. They may therefore represent attractive opportunities for therapeutic intervention, provided they are tractable for drug development. The Open Targets tractability assessment categorises genes into levels of potential tractability as drug targets by integrating information such as the clinical precedence, structural characterisation, protein family, and cellular location (Methods).

We used the Open Targets tractability assessment to explore the drug development opportunities within the TargetAge gene set (Fig 3b). Of the 995 targets in the TargetAge set, 86 have associated drugs with approved indications, and a further 42 have associated drugs currently in clinical trials. There are approved small molecule drugs for 65 targets, therapeutic antibodies for 9 targets, and 22 are targeted by therapeutics of other modalities. One target, oestrogen receptor 1 (*ESR1*), is the target of an investigational proteolysis-targeting chimeras (PROTAC) treatment for Metastatic ER+/HER2- Breast Cancer. Targets with existing drugs may have efficacy and safety information available and may represent potential drug repurposing opportunities.

For the development of a small molecule drug, targets must have a suitable binding site. There is evidence of ligand binding for 202 TargetAge targets (categorised as “Discovery Precedence”), and a further 61 targets are predicted to have a binding pocket or are members of the druggable genome (categorised as “Predicted Tractable”) (see Methods). The druggable genome represents the subset of human genes thought to be amenable to drug development, including those closely related to drug targets, secreted proteins, and members of key drug target protein families (Finan et al. 2017).

For a therapeutic antibody approach, a target is categorised as “Predicted Tractable” if its subcellular localisation is accessible to an antibody, such as location in the plasma membrane. There are 324 high confidence “Predicted Tractable” targets that are annotated to accessible locations based on high confidence experimental evidence, while 158 medium confidence targets are either annotated with medium confidence evidence or have predicted Signal Peptide or Trans-membrane regions (see Methods).

Proteolysis-targeting chimeras (PROTACs) are an emerging drug modality in which proteins are targeted for ubiquitin-directed degradation (Schneider et al. 2021). There are literature reports of successful PROTAC approaches for 28 targets, and a further 772 represent possible PROTAC opportunities based on the PROTACtable Genome assessment (Schneider et al. 2021), which considers the cellular location, evidence of ubiquitination and small-molecule ligand binding sites, and the availability of protein half-life information (see Methods).

Chemical probes are potent, selective, cell permeable compounds that modulate protein function and thereby facilitate target validation and early stage drug discovery (Arrowsmith et al. 2015; Gerry and Schreiber 2018). High quality chemical probes are available for 38 TargetAge targets, characterised by the expert-curated Chemical Probes Portal (Arrowsmith et al. 2015), the precompetitive pharmaceutical collaboration Open Science Probes (Müller et al. 2018), or the public-private partnership Structural Genomics Consortium (Arrowsmith et al. 2015). Of these, 11 targets do not have existing drugs in clinical development, representing potential novel drug discovery opportunities (SI Table 4). Furthermore, around one fifth of the TargetAge genes (n=138) have at least one potential chemical probe identified by Probe Miner, a public resource that systematically assesses and ranks small molecules for potential use as chemical probes based on public bioactivity data (Antolin et al. 2018).

Overall, 128 (13%) TargetAge genes are the target of an approved or investigational drug. A further 526 (53%) have experimental evidence of binding pockets or are predicted to be tractable by small molecule or antibody modality approaches. The remaining 341 (34%) have no existing tractability evidence, representing underexplored genes which may reveal novel biological insights and therapeutic opportunities.

### The TargetAge web application enables target exploration and triage

We developed a web application (https://targetage.shinyapps.io/TargetAge/) to facilitate exploration of the annotations of TargetAge genes identified as genetically implicated in multiple age-related traits (Fig 4).

**Fig 4.**
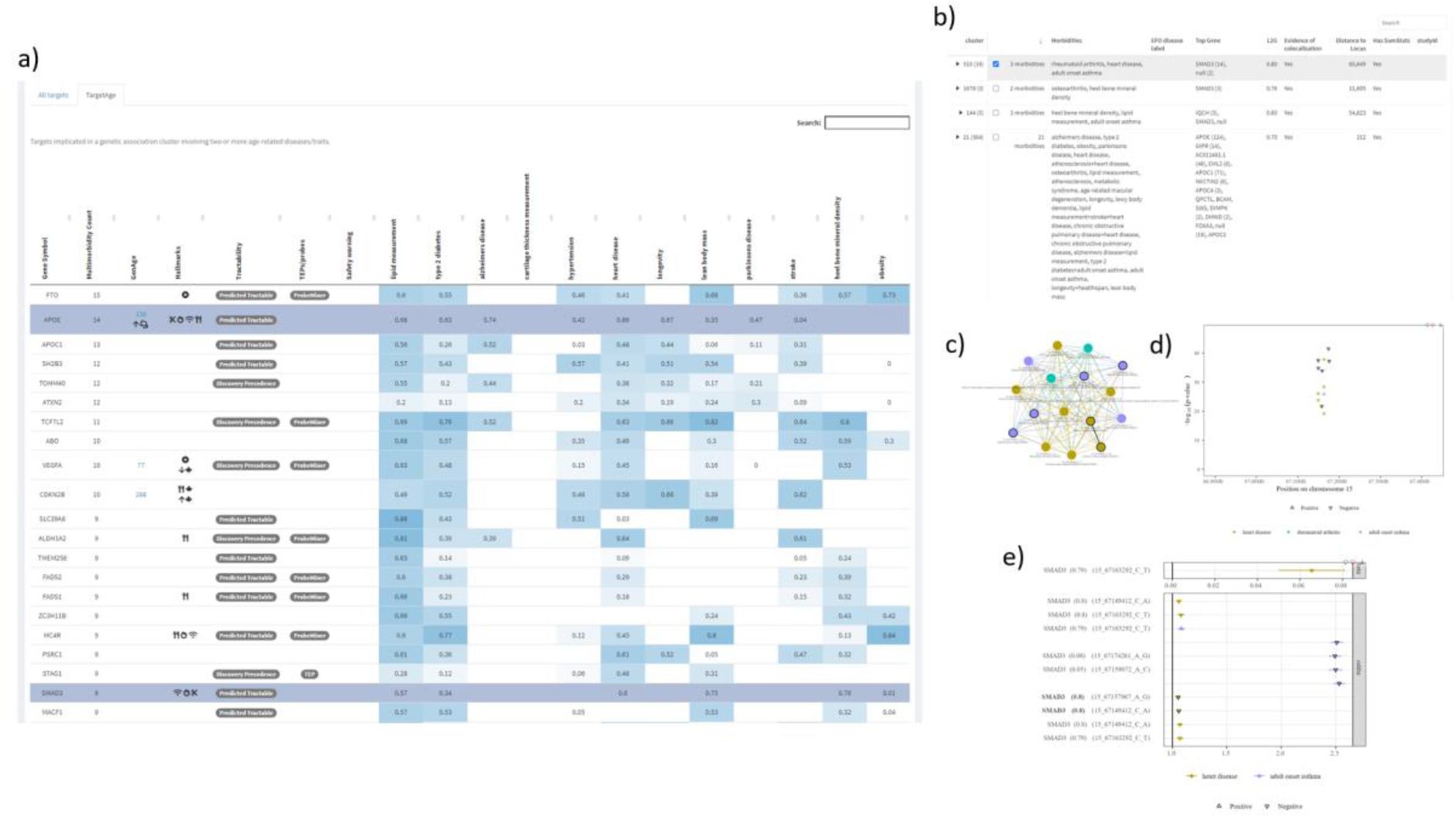
**a**) The list of TargetAge genes is shown within the TargetAge application, along with the strength of the genetic link to each age-related trait, known links to ageing or the Hallmarks of Ageing, predicted tractability, and available chemical probes or TEPs. b) Details of all multimorbidity clusters are shown, including the number of communities, the most likely causal genes, and information about the GWAS studies and lead variants associated with the cluster. c) A selected cluster can be explored using a network visualisation. d) The genomic positions and significance of lead variants associated with a cluster. e) The effect size (beta coefficient or odds ratio) of each lead variant, along with the L2G score for the most likely causal gene.

Each target is annotated with the strength of the genetic link to each age-related trait (the Open Targets genetic association score), known links to ageing (GenAge, CellAge, and Hallmarks of Ageing), as well as the clinical precedence, tractability assessment, and known safety flags. The results can be filtered for specific genes, age-related traits, or Hallmarks of Ageing. In addition, individual genetic association clusters can be visualised and explored. Information on each study and locus in the cluster is presented, including the trait studied, sample size, effect size, most likely causal gene and L2G score. For cluster visualisation in TargetAge, nodes with summary statistics are highlighted with a black outline, edges supported by co-localisation are indicated with a thick black line, and pairs of nodes that share LD-defined tag variants but are not likely to share a causal variant according to co-localisation analysis are indicated with a dashed line.

## Discussion

We systematically combined 1,394 GWAS of age-related traits to identify clusters where one or more of these traits shared a common genetic signal. To develop the genetic clusters, we made use of genetic co-localisation (when summary statistics were available and credible sets overlapped) and LD-expansion (in the absence of summary statistics) from Open Targets Genetics. We identified 1,006 clusters in which a shared causal variant is implicated in multiple age-related traits. From these clusters, we created the TargetAge gene set of 995 candidate causal genes, of which 29% have a known annotation to at least one Hallmark of Ageing. This set has a statistically significant overlap with the manually curated ageing-related gene sets GenAge and CellAge, with evidence of shared biological processes, but 935 of these genes are novel.

The TargetAge genes and genetic analysis outputs are presented in an interactive web application. The multimorbidity genetic clusters and candidate genes are displayed alongside evidence of links to ageing and predicted tractability information. Individual comorbidity clusters reveal potentially shared genetic signals between multiple age-related traits and offer further avenues for investigation. Where summary statistics are available, co-localisation evidence indicates where associations have the same direction of effect.

The application facilitates the exploration and prioritisation of potential drug development opportunities for age-related multimorbidity. Some may represent drug repurposing opportunities: 13% of TargetAge genes are targeted by drugs with investigational or approved indications. A further 18% have experimental evidence of binding pockets, and a further 34% are predicted tractable either by small molecule or antibody therapeutics. Of these, 11 targets have high quality chemical probes already available and no existing drugs in clinical development. The remaining 34% have no existing tractability evidence, representing underexplored genes which may reveal novel biological insights and therapeutic opportunities. Together this analysis suggests there is substantial underexploited opportunity for development of drugs for age-related multi-morbidities.

Some clusters identified in this analysis reflect established relationships between commonly occurring comorbidities. For example, a locus associated with atrial fibrillation and heel bone mineral density is linked to thyroid hormone receptor beta (*THRB*) - one of several receptors for thyroid hormones - in cluster 1307. Overstimulation of this receptor by thyroid hormone can cause hyperthyroidism (Brent 2012). In adults, hyperthyroidism is associated with increased incidence of atrial fibrillation - with older patients at greater risk (Agner et al. 1984) - as well as osteoporosis and increased risk of bone fracture, although the underlying mechanism is not fully elucidated (Delitala, Scuteri, and Doria 2020). The combination of atrial fibrillation and heel bone mineral density is represented in a further 14 clusters.

We note several clusters of cardiovascular diseases linked to a common therapeutically actionable target. For instance, cluster 209 represents an important locus for cardiovascular diseases, including coronary artery disease, peripheral arterial disease, and large artery stroke, and coronary atherosclerosis. The locus is strongly linked to the gene encoding endothelin receptor type A (*EDNRA*), the target of several approved small molecule drugs for pulmonary artery hypertension and type 2 diabetes indications, suggesting a potential repositioning opportunity for other cardiovascular diseases. Another cluster involving vascular diseases and stroke is linked to phospholipid phosphatase 3 (*PLPP3*). This gene is thought to play a role in vascular homeostasis through negative regulation of pro-inflammatory cytokines, leucocyte adhesion, cell survival and migration in aortic endothelial cells (Touat-Hamici et al. 2016). This locus co-localises with eQTLs in monocytes, which is consistent with a functional role in inflammation and vascular disease, and *PLPP3* is predicted tractable by an antibody therapeutic approach.

In some cases, shared genetic associations may represent distal regulators of the causal gene. Cluster 644 involves a common non-coding variant rs9349379 in the third intron of phosphatase and actin regulatory protein 1 (*PHACTR1*). A study by Gupta *et al*. identified this as the causal variant for associations with five vascular diseases, with a putative mechanism acting through distal regulatory effects on endothelin-1 (*EDN1*), a gene located 600 kb upstream of *PHACTR1* (Gupta et al. 2017). TargetAge replicates this association: cluster 644 links this variant to coronary artery disease, myocardial infarction, intermediate coronary syndrome, and coronary atherosclerosis. Furthermore, *PHACTR1* is implicated in a second cluster, which is associated with ischemic stroke, large artery stroke, and hand grip strength in addition to coronary artery disease and myocardial infarction.

We also find several clusters with shared therapeutically tractable targets linking measures of adiposity (e.g., body mass index) with type 2 diabetes (T2DM) risk, metabolic syndrome, and osteoarthritis. For example, *SEC16B* – a protein expressed mostly in the small intestine that is predicted tractable by antibody therapeutics – links obesity with T2DM and metabolic syndrome. *SEC16B* contributes to the intracellular trafficking of vesicles containing lipids and proteins from the endoplasmic reticulum to the Golgi apparatus for further processing and secretion (Budnik, Heesom, and Stephens 2011). A recent experimental study showed that intestinal *SEC16B* knockout mice had a significantly lower serum triglycerides after a high-fat meal, improved glucose clearance, and were protected from high fat diet-induced obesity (Shi et al. 2021). Our study, along with this experimental evidence, therefore, supports prioritization of intestinal *SEC16B* as a therapeutic target for obesity and impaired glucose tolerance.

Another example of a shared target is Filamin A interacting protein (*FILIP1*), implicated in a locus associated with increased lean body mass and increased risk of osteoarthritis (cluster 220), which is highly expressed in heart and skeletal muscle tissue, and is predicted tractable by a therapeutic antibody. We also identified shared targets that were not predicted tractable via small molecular or antibody approaches, but potentially tractable by proteolysis-targeting chimeras (PROTACs) - an emerging drug modality (Schneider et al. 2021). An example of the latter is *CMIP* (c-Maf inducing protein) - a negative regulator of T-cell signalling (Oniszczuk et al. 2020). *CMIP* is linked to cluster 57, associated with lower triglycerides, higher adiponectin (which was inversely associated with T2DM in an observational study (Li et al. 2009)), lower risk of T2DM, and lower risk of metabolic syndrome.

Our approach is highly inclusive with the aim of harnessing as much information as possible to uncover potentially novel relationships between ageing-related traits. Some of the proposed multimorbidity clusters are likely to include genetic signals that overlap but have distinct causal mechanisms, particularly in regions of low recombination and high LD, or regions with many genetic associations. Studies with full summary statistics can help discriminate distinct genetic signals that exist within a single cluster by reducing the number of possible causal variants (fine mapping) and removing connections between loci that are unlikely to share a causal variant despite sharing some overlapping tag variants (co-localisation). However, studies with summary statistics represent only around 10% of the GWAS studies available for the investigated ageing-related traits. Increased availability of comprehensive summary statistics would therefore improve our ability to distinguish overlapping but distinct signals. Community detection and visual inspection can help evaluate whether an individual cluster represents a shared genetic signal; the presence of multiple communities within the cluster may suggest that it includes distinct genetic signals.

The methodology described here demonstrates one way in which large-scale tools such as Open Targets Platform and Open Targets Genetics can be used to systematically combine evidence from difference sources and use co-localisation data at scale for multiple diseases and traits. Rather than understanding and treating ageing-related diseases seen in multimorbid patients in isolation, there is growing evidence that better health outcomes may be achieved by recognising that diseases often co-occur non-randomly around common genetic, biological, or environmental pathways (MacMahon et al. 2018; Whitty et al. 2020). Such Geroscience, multimorbidity focussed approaches are relevant beyond age-related diseases, and the methodology described here could be applied to other clinically relevant combinations of diseases thought to share common genetic or biological pathways.

We have shown that our approach is able to highlight a range of opportunities both in terms of existing therapeutic agents against linked targets and also novel targets which might allow treatment for multimorbidity. It will be important to develop tool molecules for those novel targets identified in order to better understand their biology and to validate their possible role in disease. Ultimately, clinical trials will be required to fully validate the associated proposed hypotheses in an ageing and multimorbidity context.

## Methods

### Genetic Analysis

#### Data

Target and GWAS data were obtained from Open Targets Platform version 22.02 and Open Targets Genetics version 210608. From Open Targets Platform, targets were retrieved for which there is GWAS evidence linking the gene to any of the age-related trait EFO codes, or their descendant terms in the Open Targets EFO slim, excluding terms that contain the word “juvenile” or fall under the therapeutic areas “genetic, familial or congenital disease” (OTAR_0000018), “pregnancy or perinatal disease” (OTAR_0000014), or “cell proliferation disorder” (MONDO_0045024). For all the GWAS studies contributing to this evidence, study information and lead variants were retrieved from Open Targets Genetics.

#### Lead to tag variant expansion

The lead variant represents just one possible causal variant for each genetic association. These are expanded to a set of possible causal variants, referred to as tag variants. Open Targets Genetics uses two methods to perform this expansion: Linkage Disequilibrium (LD) expansion and fine-mapping expansion.

#### Linkage Disequilibrium (LD) expansion

Where summary statistics are not available, the tag variants include all those in LD with the lead variant (r^2^>=0.7). In Open Targets Genetics, LD is calculated for all studies using the 1000 Genomes Phase 3 (1KG) genotypes as a reference. Where ancestry information is known for the GWAS study population, the most closely matching 1KG super-population is used - or a weighted-average across relevant super-populations for mixed ancestry study populations - otherwise European ancestry is assumed.

#### Fine mapping expansion

Details of fine-mapping conducted for Open Targets Genetics are provided elsewhere (Mountjoy et al. 2021; ‘Assigning Variants to Disease (V2D)’ n.d.). In brief, where summary statistics were available, Genome-wide Complex Trait Analysis Conditional and Joint Analysis (GCTA-COJO; v1.91.3 (Yang et al. 2012)) was used to identify conditionally independent loci (where both the marginal and conditional p-values were less than 5 × 10^−8^) using genotypes from the UK Biobank population down-sampled to 10K study participants as the linkage-disequilibrium (LD) reference for the conditional analysis. Approximate Bayes factors were computed using the conditional summary statistics and posterior probabilities (PP) were derived from the Bayes factors for all SNPs within a ±500Kb window assuming a single causal variant. Where summary statistics were not available, we used the Probabilistic Identification of Causal SNPs (PICS) method (Farh et al. 2015) with LD from 1000 Genomes phase 3 data to estimate the PP for each variant. In both cases, any variant with a PP > 0.1% in the credible set is retained.

#### GWAS association overlap

We used two methods to determine whether associations from different GWAS studies may share a causal variant and therefore represent the same signal, depending on the availability of full summary statistics for those studies.

Where summary statistics are available, Open Targets Genetics co-localisation analysis is used (Ghoussaini et al. 2021; Giambartolomei et al. 2014). For each locus, this method integrates over evidence from all variants within a ±500Kb window in each study to evaluate which of these four hypotheses is most likely: no association with either trait (H0), association only with trait 1 (H1), association only with trait 2 (H2), association with both traits via two independent SNPs (H3), or association with both traits through a shared causal SNP (H4). We use a cut-off of H4>=0.8 to define the association for study 1 and study 2 as sharing the same causal variant.

For most cases, summary statistics are not available for one or both studies. In this case, for lead variants less than 500Kb apart, the associations are considered to represent the same signal if at least one of the tag variants is shared between the two lead variants. Using a more stringent overlap criterion increased the overall number of clusters and decreased the number of clusters involving more than one age-related trait, but does not substantially affect the structure of the graph (SI Fig 1a). When considering the number of communities detected within each cluster, more stringent criteria reduced the outliers with a high number of communities but did not substantially affect the overall distribution, suggesting the clusters are relatively robust to this criterion (SI Fig 1b). For inclusivity, we use the least stringent definition of an overlap.

#### Clustering genetic associations

We construct an undirected, unweighted graph in which each node is a trait-associated locus from a single GWAS study, represented by the GWAS study and the lead variant. Nodes are connected by an edge where these genetic associations may share a causal variant, either through co-localisation analysis (where summary statistics are available) or tag variant overlap (see *GWAS association overlap*, above).

Graph construction and analysis was carried out in R (version 4.0.2) using igraph (version 1.2.6) (Csardi and Nepusz 2006) and visnetwork (version 2.0.9) (Almende B.V., Thieurmel, and Robert 2019) for graph visualisation. Disconnected clusters within the graph represent independent genetic signals, and are identified by calculating the maximal connected components, using the “clusters” function (also known as “components”) with default parameters. To identify substructure within each cluster, which may indicate separate signals, we identify communities using “cluster_louvain” implementation of the Louvain community detection method with default parameters (Blondel et al. 2008).

To test the statistical significance of the genetic overlap between each combination of traits, raw p-values were obtained using one-sided Fishers exact test for each pair, implemented using *fisher*.*exact* in R. P-values adjusted for multiple testing (406 possible combinations of 29 traits) were obtained using BH (also known as FDR) correction implemented in the *p*.*adjust* function (Benjamini and Hochberg 1995).

#### Assigning causal genes to GWAS loci

Loci were assigned to likely causal genes using the Open Targets Genetics locus-to-gene (L2G) score (Mountjoy et al. 2021). This score ranges from 0 to 1 and is assigned using a supervised machine learning model, which is trained on predictive functional genomics features (such as predicted pathogenicity and genomic distance) for 400 gold standard GWAS loci with high-confidence causal genes. We only consider genes with an L2G score of at least 0.5 and assign each locus to the gene with the highest L2G score.

### Ageing-related gene sets

GenAge is a manually curated database of genes thought to be involved in ageing or longevity that includes 307 human genes (GenAge Human) and 2,205 genes from model organisms (GenAge Models) (Tacutu et al. 2018). Human genes are included in the database if there is experimental evidence linking the gene to ageing through gene manipulation experiments in human, mammal, or animal models.

CellAge is a manually-curated database of 279 genes found to induce or inhibit cellular senescence through gene manipulation experiments in human cell models (Avelar et al. 2020). Cellular senescence is one of the pathways associated with ageing and age-related pathology, and a major target for therapeutic intervention (Amaya-Montoya et al. 2020).

The Hallmarks of Ageing are processes that manifest during normal ageing, for which experimental manipulation can accelerate or decelerate the ageing process (López-Otín et al. 2013). This includes causes of cellular damage (genomic instability, telomere attrition, epigenetic alterations, and loss of proteostasis), responses to damage (deregulated nutrient sensing, mitochondrial dysfunction, and cellular senescence), and processes that lead to the ageing trait (stem cell exhaustion, altered intracellular communication). We curated a subset of Gene Ontology (GO) biological process terms that fall under each of the hallmarks of ageing (SI Table 3).

### Tractability assessment

Target tractability assessment information was retrieved from Open Targets, which uses a pipeline based on (Brown et al. 2018) and PROTAC tractability workflow from (Schneider et al. 2021).

Targets are classified as “Clinical Precedence” if there are drugs approved or in clinical development based on curated mechanisms of action from ChEMBL (Mendez et al. 2019).

For small molecule modality, targets are categorised as “Discovery Precedence” if there is evidence of ligand binding (either high quality compounds with bioactivity data in ChEMBL or co-crystallisation with a ligand) and “Predicted Tractable” if the target is predicted to have a binding pocket (drugEBIlity score >= 0.7) and/or is a member of the druggable genome.

For antibody modality, targets categorised as “Predicted Tractable high confidence” are localised to the plasma membrane, extracellular matrix, or secreted, according to high confidence “Subcellular location” terms in UniProt or “Cellular component” terms in GO. “Predicted Tractable Medium to low confidence” targets are annotated to these locations with medium confidence or have predicted Signal Peptide or Trans-membrane regions.

For PROTAC modality, targets with literature reports of successful PROTAC degrader are classified as “Literature Precedence”. Targets are classified as “Discovery Opportunity” if they meet the following criteria: evidence of a ubiquitylation site (UniProt, PhosphoSitePlus, mUbiSiDa, or diglycine proteomics experiments); available experimental data on protein half-life; small-molecule ligand binders (≥10uM activity reported in ChEMBL), are secreted, extracellular, or located in the cell cytoplasm, cytosol or nucleus (“Subcellular location” terms in UniProt or “Cellular component” terms in GO).

#### Curated chemical probes

Information on available high quality chemical probes is retrieved through Open Targets. Chemical probes released by the Structural Genomics Consortium (https://www.thesgc.org/chemical-probes) and through Open Science Probes (https://www.sgc-ffm.uni-frankfurt.de/) are open access with no restrictions on use and are required to meet the following criteria: *in vitro* potency of <100 nm, >30-fold selectivity compared to related proteins within the same target family, demonstrated on-target effects in cells at <1 μM, no PAINS elements, and (more recently) an inactive close analogue suitable for use as a control (Arrowsmith et al. 2015). The Chemical Probes Portal (https://www.chemicalprobes.org/) provides expert guidance and ratings for chemical probes; probes are included in Open Targets if they receive a rating of 4 or 3 stars, indicating a recommended or best available probe for a target.

#### Predicted chemical probes

Probe Miner ranks all bioactive compounds for a particular protein target based on six scores: potency, selectivity, cellular activity (as a proxy for cell permeability), structure-activity relationships (SAR), the availability of inactive analogues, and pan-assay interference compound (PAINS) prediction (Antolin et al. 2018).

## Data Availability

All data used in the study is publicly available, and all code and data produced in the study is available at https://github.com/CMD-Oxford/targetage-pipeline and https://github.com/CMD-Oxford/TargetAgeApp

https://genetics.opentargets.org/

https://github.com/CMD-Oxford/targetage-pipeline

## Code availability

Code for the analysis and the web applications is available on GitHub at https://github.com/CMD-Oxford/targetage-pipeline and https://github.com/CMD-Oxford/TargetAgeApp.

## Acknowledgements

C.E.W., B.V., J.M.L., G.W., C.J.G. and L.L. are funded by the UK SPINE knowledge exchange network. B.M. is funded by the Kennedy Trust for Rheumatology Research. L.S. Acknowledges the support provided by the Sir Henry Wellcome fellowship [220457/Z/20/z]. M.K., M.J.F., J.S., D.O. and I.D. are funded by Open Targets. This research was funded in part by a Wellcome Trust [Grant number 206194]. For the purpose of Open Access, the authors have applied a CC-BY public copyright licence to any author-accepted manuscript version arising from this submission.

## Contributions

C.E.W. wrote the manuscript with contributions from B.D.M., I.D., A.R.L., M.K., L.S., J.M.L. and J.S‥ C.E.W. designed and performed the analysis with the help of M.K., L.S., J.S., D.O. and M.J.F. and D.O., and interpreted the results with contributions from all authors. B.D.M, I.D., A.R.L., B.V., and C.B. conceived and supervised the study.

## References

Agner, T., T. Almdal, B. Thorsteinsson, and E. Agner. 1984. ‘A Reevaluation of Atrial Fibrillation in Thyrotoxicosis’. Danish Medical Bulletin 31 (2): 157–59. https://www.ncbi.nlm.nih.gov/pubmed/6723378.

Almende B.V., Benoit Thieurmel, and Titouan Robert. 2019. ‘VisNetwork: Network Visualization Using “vis.Js” Library’. https://CRAN.R-project.org/package=visNetwork.

Amaya-Montoya, Mateo, Agustín Pérez-Londoño, Valentina Guatibonza-García, Andrea Vargas-Villanueva, and Carlos O. Mendivil. 2020. ‘Cellular Senescence as a Therapeutic Target for Age-Related Diseases: A Review’. Advances in Therapy 37 (4): 1407–24. https://doi.org/10.1007/s12325-020-01287-0.

Antolin, Albert A., Joseph E. Tym, Angeliki Komianou, Ian Collins, Paul Workman, and Bissan Al-Lazikani. 2018. ‘Objective, Quantitative, Data-Driven Assessment of Chemical Probes’. Cell Chemical Biology 25 (2): 194-205.e5. https://doi.org/10.1016/j.chembiol.2017.11.004.

Argmann, Carmen, Radu Dobrin, Sami Heikkinen, Aurélie Auburtin, Laurent Pouilly, Terrie-Anne Cock, Hana Koutnikova, Jun Zhu, Eric E. Schadt, and Johan Auwerx. 2009. ‘Ppargamma2 Is a Key Driver of Longevity in the Mouse’. PLoS Genetics 5 (12): e1000752. https://doi.org/10.1371/journal.pgen.1000752.

Arrowsmith, Cheryl H., James E. Audia, Christopher Austin, Jonathan Baell, Jonathan Bennett, Julian Blagg, Chas Bountra, et al. 2015. ‘The Promise and Peril of Chemical Probes’. Nature Chemical Biology 11 (8): 536–41. https://doi.org/10.1038/nchembio.1867.

‘Assigning Variants to Disease (V2D)’. n.d. Accessed 17 April 2022. https://genetics-docs.opentargets.org/our-approach/assigning-traits-to-loci.

Avelar, Roberto A., Javier Gómez Ortega, Robi Tacutu, Eleanor J. Tyler, Dominic Bennett, Paolo Binetti, Arie Budovsky, et al. 2020. ‘A Multidimensional Systems Biology Analysis of Cellular Senescence in Aging and Disease’. Genome Biology 21 (1): 91. https://doi.org/10.1186/s13059-020-01990-9.

Barzilai, Nir, Jill P. Crandall, Stephen B. Kritchevsky, and Mark A. Espeland. 2016. ‘Metformin as a Tool to Target Aging’. Cell Metabolism 23 (6): 1060–65. https://doi.org/10.1016/j.cmet.2016.05.011.

Belloy, Michaël E., Valerio Napolioni, and Michael D. Greicius. 2019. ‘A Quarter Century of APOE and Alzheimer’s Disease: Progress to Date and the Path Forward’. Neuron 101 (5): 820–38. https://doi.org/10.1016/j.neuron.2019.01.056.

Benjamini, Yoav, and Yosef Hochberg. 1995. ‘Controlling the False Discovery Rate: A Practical and Powerful Approach to Multiple Testing’. Journal of the Royal Statistical Society 57 (1): 289–300. https://doi.org/10.1111/j.2517-6161.1995.tb02031.x.

Blondel, Vincent D., Jean-Loup Guillaume, Renaud Lambiotte, and Etienne Lefebvre. 2008. ‘Fast Unfolding of Communities in Large Networks’. Journal of Statistical Mechanics 2008 (10): P10008. https://doi.org/10.1088/1742-5468/2008/10/P10008.

Bohannon, Richard W. 2019. ‘Grip Strength: An Indispensable Biomarker For Older Adults’. Clinical Interventions in Aging 14 (October): 1681–91. https://doi.org/10.2147/CIA.S194543.

Brent, Gregory A. 2012. ‘Mechanisms of Thyroid Hormone Action’. The Journal of Clinical Investigation 122 (9): 3035–43. https://doi.org/10.1172/JCI60047.

Brown, Kristin K., Michael M. Hann, Ami S. Lakdawala, Rita Santos, Pamela J. Thomas, and Kieran Todd. 2018. ‘Approaches to Target Tractability Assessment - a Practical Perspective’. MedChemComm 9 (4): 606–13. https://doi.org/10.1039/c7md00633k.

Budnik, Annika, Kate J. Heesom, and David J. Stephens. 2011. ‘Characterization of Human Sec16B: Indications of Specialized, Non-Redundant Functions’. Scientific Reports 1 (August): 77. https://doi.org/10.1038/srep00077.

Calderón-Larrañaga, A., D. L. Vetrano, L. Ferrucci, S. W. Mercer, A. Marengoni, G. Onder, M. Eriksdotter, and L. Fratiglioni. 2019. ‘Multimorbidity and Functional Impairment-Bidirectional Interplay, Synergistic Effects and Common Pathways’. Journal of Internal Medicine 285 (3): 255–71. https://doi.org/10.1111/joim.12843.

Chaib, Selim, Tamar Tchkonia, and James L. Kirkland. 2022. ‘Cellular Senescence and Senolytics: The Path to the Clinic’. Nature Medicine 28 (8): 1556–68. https://doi.org/10.1038/s41591-022-01923-y.

Clancy, D. J., D. Gems, L. G. Harshman, S. Oldham, H. Stocker, E. Hafen, S. J. Leevers, and L. Partridge. 2001. ‘Extension of Life-Span by Loss of CHICO, a Drosophila Insulin Receptor Substrate Protein’. Science 292 (5514): 104–6. https://doi.org/10.1126/science.1057991.

Cruz-Jentoft, Alfonso J., Gülistan Bahat, Jürgen Bauer, Yves Boirie, Olivier Bruyère, Tommy Cederholm, Cyrus Cooper, et al. 2019. ‘Sarcopenia: Revised European Consensus on Definition and Diagnosis’. Age and Ageing 48 (1): 16–31. https://doi.org/10.1093/ageing/afy169.

Dai, Haiming, X. Wei Meng, Kaiqin Ye, Jia Jia, and Scott H. Kaufmann. 2022. ‘Chapter 7 - Therapeutics Targeting BCL2 Family Proteins’. In Mechanisms of Cell Death and Opportunities for Therapeutic Development, edited by Daiqing Liao, 197–260. Academic Press. https://doi.org/10.1016/B978-0-12-814208-0.00007-5.

Delitala, Alessandro P., Angelo Scuteri, and Carlo Doria. 2020. ‘Thyroid Hormone Diseases and Osteoporosis’. Journal of Clinical Medicine Research 9 (4). https://doi.org/10.3390/jcm9041034.

Dönertaş, Handan Melike, Daniel K. Fabian, Matías Fuentealba, Linda Partridge, and Janet M. Thornton. 2021. ‘Common Genetic Associations between Age-Related Diseases’. Nature Aging, April, 1–13. https://doi.org/10.1038/s43587-021-00051-5.

Dönertaş, Handan Melike, Matías Fuentealba Valenzuela, Linda Partridge, and Janet M. Thornton. 2018. ‘Gene Expression-Based Drug Repurposing to Target Aging’. Aging Cell 17 (5): e12819. https://doi.org/10.1111/acel.12819.

Ermogenous, Christos, Charlotte Green, Thomas Jackson, Michael Ferguson, and Janet M. Lord. 2020. ‘Treating Age-Related Multimorbidity: The Drug Discovery Challenge’. Drug Discovery Today 25 (8): 1403–15. https://doi.org/10.1016/j.drudis.2020.06.016.

Farh, Kyle Kai-How, Alexander Marson, Jiang Zhu, Markus Kleinewietfeld, William J. Housley, Samantha Beik, Noam Shoresh, et al. 2015. ‘Genetic and Epigenetic Fine Mapping of Causal Autoimmune Disease Variants’. Nature 518 (7539): 337–43. https://doi.org/10.1038/nature13835.

Finan, Chris, Anna Gaulton, Felix A. Kruger, R. Thomas Lumbers, Tina Shah, Jorgen Engmann, Luana Galver, et al. 2017. ‘The Druggable Genome and Support for Target Identification and Validation in Drug Development’. Science Translational Medicine 9 (383). https://doi.org/10.1126/scitranslmed.aag1166.

Gerry, Christopher J., and Stuart L. Schreiber. 2018. ‘Chemical Probes and Drug Leads from Advances in Synthetic Planning and Methodology’. Nature Reviews. Drug Discovery 17 (5): 333–52. https://doi.org/10.1038/nrd.2018.53.

Geyer, Orna, and Yoram Levo. 2020. ‘Glaucoma Is an Autoimmune Disease’. Autoimmunity Reviews 19 (6): 102535. https://doi.org/10.1016/j.autrev.2020.102535.

Ghoussaini, Maya, Edward Mountjoy, Miguel Carmona, Gareth Peat, Ellen M. Schmidt, Andrew Hercules, Luca Fumis, et al. 2021. ‘Open Targets Genetics: Systematic Identification of Trait-Associated Genes Using Large-Scale Genetics and Functional Genomics’. Nucleic Acids Research 49 (D1): D1311–20. https://doi.org/10.1093/nar/gkaa840.

Giambartolomei, Claudia, Damjan Vukcevic, Eric E. Schadt, Lude Franke, Aroon D. Hingorani, Chris Wallace, and Vincent Plagnol. 2014. ‘Bayesian Test for Colocalisation between Pairs of Genetic Association Studies Using Summary Statistics’. PLoS Genetics 10 (5): e1004383. https://doi.org/10.1371/journal.pgen.1004383.

Gupta, Rajat M., Joseph Hadaya, Aditi Trehan, Seyedeh M. Zekavat, Carolina Roselli, Derek Klarin, Connor A. Emdin, et al. 2017. ‘A Genetic Variant Associated with Five Vascular Diseases Is a Distal Regulator of Endothelin-1 Gene Expression’. Cell 170 (3): 522-533.e15. https://doi.org/10.1016/j.cell.2017.06.049.

Kim, Seung Hoon, Sung Hoon Jeong, Hyunkyu Kim, Eun-Cheol Park, and Suk-Yong Jang. 2022. ‘Development of Open-Angle Glaucoma in Adults With Seropositive Rheumatoid Arthritis in Korea’. JAMA Network Open 5 (3): e223345. https://doi.org/10.1001/jamanetworkopen.2022.3345.

Kim, So Young, Chanyang Min, Dong Jun Oh, and Hyo Geun Choi. 2019. ‘Increased Risk of Asthma in Patients with Rheumatoid Arthritis: A Longitudinal Follow-up Study Using a National Sample Cohort’. Scientific Reports 9 (1): 6957. https://doi.org/10.1038/s41598-019-43481-3.

King, Emily A., J. Wade Davis, and Jacob F. Degner. 2019. ‘Are Drug Targets with Genetic Support Twice as Likely to Be Approved? Revised Estimates of the Impact of Genetic Support for Drug Mechanisms on the Probability of Drug Approval’. PLoS Genetics 15 (12): e1008489. https://doi.org/10.1371/journal.pgen.1008489.

Kumar P, Pavan, Uchenna Emechebe, Richard Smith, Sarah Franklin, Barry Moore, Mark Yandell, Stephen L. Lessnick, and Anne M. Moon. 2014. ‘Coordinated Control of Senescence by LncRNA and a Novel T-Box3 Co-Repressor Complex’. ELife 3 (May). https://doi.org/10.7554/eLife.02805.

Li, Shanshan, Hyun Joon Shin, Eric L. Ding, and Rob M. van Dam. 2009. ‘Adiponectin Levels and Risk of Type 2 Diabetes: A Systematic Review and Meta-Analysis’. JAMA: The Journal of the American Medical Association 302 (2): 179–88. https://doi.org/10.1001/jama.2009.976.

Liu, Ying, Lijia Yu, Yaling Xu, Xiaohui Tang, and Xijin Wang. 2020. ‘Substantia Nigra Smad3 Signaling Deficiency: Relevance to Aging and Parkinson’s Disease and Roles of Microglia, Proinflammatory Factors, and MAPK’. Journal of Neuroinflammation 17 (1): 342. https://doi.org/10.1186/s12974-020-02023-9.

López-Otín, Carlos, Maria A. Blasco, Linda Partridge, Manuel Serrano, and Guido Kroemer. 2013. ‘The Hallmarks of Aging’. Cell 153 (6): 1194–1217. https://doi.org/10.1016/j.cell.2013.05.039.

MacMahon, S., P. Calverley, N. Chaturvedi, Z. Chen, L. Corner, M. Davies, M. Ezzati, et al. 2018. ‘Multimorbidity: A Priority for Global Health Research’. The Academy of Medical Sciences: London, UK, 127.

Malone, James, Ele Holloway, Tomasz Adamusiak, Misha Kapushesky, Jie Zheng, Nikolay Kolesnikov, Anna Zhukova, Alvis Brazma, and Helen Parkinson. 2010. ‘Modeling Sample Variables with an Experimental Factor Ontology’. Bioinformatics 26 (8): 1112–18. https://doi.org/10.1093/bioinformatics/btq099.

Mendez, David, Anna Gaulton, A. Patrícia Bento, Jon Chambers, Marleen De Veij, Eloy Félix, María Paula Magariños, et al. 2019. ‘ChEMBL: Towards Direct Deposition of Bioassay Data’. Nucleic Acids Research 47 (D1): D930–40. https://doi.org/10.1093/nar/gky1075.

Mountjoy, Edward, Ellen M. Schmidt, Miguel Carmona, Jeremy Schwartzentruber, Gareth Peat, Alfredo Miranda, Luca Fumis, et al. 2021. ‘An Open Approach to Systematically Prioritize Causal Variants and Genes at All Published Human GWAS Trait-Associated Loci’. Nature Genetics 53 (11): 1527–33. https://doi.org/10.1038/s41588-021-00945-5.

Müller, Susanne, Suzanne Ackloo, Cheryl H. Arrowsmith, Marcus Bauser, Jeremy L. Baryza, Julian Blagg, Jark Böttcher, et al. 2018. ‘Donated Chemical Probes for Open Science’. ELife 7 (April). https://doi.org/10.7554/eLife.34311.

Mutlu, Ayse Sena, Jonathon Duffy, and Meng C. Wang. 2021. ‘Lipid Metabolism and Lipid Signals in Aging and Longevity’. Developmental Cell 56 (10): 1394–1407. https://doi.org/10.1016/j.devcel.2021.03.034.

Ochoa, David, Andrew Hercules, Miguel Carmona, Daniel Suveges, Asier Gonzalez-Uriarte, Cinzia Malangone, Alfredo Miranda, et al. 2020. ‘Open Targets Platform: Supporting Systematic Drug–Target Identification and Prioritisation’. Nucleic Acids Research 49 (D1): D1302–10. https://doi.org/10.1093/nar/gkaa1027.

Oniszczuk, Julie, Kelhia Sendeyo, Cerina Chhuon, Berkan Savas, Etienne Cogné, Pauline Vachin, Carole Henique, et al. 2020. ‘CMIP Is a Negative Regulator of T Cell Signaling’. Cellular & Molecular Immunology 17 (10): 1026–41. https://doi.org/10.1038/s41423-019-0266-5.

Partridge, Linda, Joris Deelen, and P. Eline Slagboom. 2018. ‘Facing up to the Global Challenges of Ageing’. Nature 561 (7721): 45–56. https://doi.org/10.1038/s41586-018-0457-8.

Pun, Frank W., Geoffrey Ho Duen Leung, Hoi Wing Leung, Bonnie Hei Man Liu, Xi Long, Ivan V. Ozerov, Ju Wang, et al. 2022. ‘Hallmarks of Aging-Based Dual-Purpose Disease and Age-Associated Targets Predicted Using PandaOmics AI-Powered Discovery Engine’. Aging 14 (6): 2475–2506. https://doi.org/10.18632/aging.203960.

Rochlani, Yogita, Naga Venkata Pothineni, Swathi Kovelamudi, and Jawahar L. Mehta. 2017. ‘Metabolic Syndrome: Pathophysiology, Management, and Modulation by Natural Compounds’. Therapeutic Advances in Cardiovascular Disease 11 (8): 215–25. https://doi.org/10.1177/1753944717711379.

Rolfes, Mary Claire, Young Jun Juhn, Chung-Il Wi, and Youn Ho Sheen. 2017. ‘Asthma and the Risk of Rheumatoid Arthritis: An Insight into the Heterogeneity and Phenotypes of Asthma’. Tuberculosis and Respiratory Diseases 80 (2): 113–35. https://doi.org/10.4046/trd.2017.80.2.113.

Schimunek, Lukas, Rami A. Namas, Jinling Yin, Derek Barclay, Dongmei Liu, Fayten El-Dehaibi, Andrew Abboud, et al. 2019. ‘MPPED2 Polymorphism Is Associated With Altered Systemic Inflammation and Adverse Trauma Outcomes’. Frontiers in Genetics 10 (November): 1115. https://doi.org/10.3389/fgene.2019.01115.

Schneider, Melanie, Chris J. Radoux, Andrew Hercules, David Ochoa, Ian Dunham, Lykourgos-Panagiotis Zalmas, Gerhard Hessler, et al. 2021. ‘The PROTACtable Genome’. Nature Reviews. Drug Discovery 20 (10): 789–97. https://doi.org/10.1038/s41573-021-00245-x.

Selman, Colin, Steven Lingard, Agharul I. Choudhury, Rachel L. Batterham, Marc Claret, Melanie Clements, Faruk Ramadani, et al. 2008. ‘Evidence for Lifespan Extension and Delayed Age-Related Biomarkers in Insulin Receptor Substrate 1 Null Mice’. FASEB Journal: Official Publication of the Federation of American Societies for Experimental Biology 22 (3): 807–18. https://doi.org/10.1096/fj.07-9261com.

Shi, Ruicheng, Wei Lu, Ye Tian, and Bo Wang. 2021. ‘Intestinal SEC16B Modulates Obesity by Controlling Dietary Lipid Absorption’. BioRxiv. https://doi.org/10.1101/2021.12.07.471468.

Sun, Dianjianyi, Tao Zhou, Yoriko Heianza, Xiang Li, Mengyu Fan, Vivian A. Fonseca, and Lu Qi. 2019. ‘Type 2 Diabetes and Hypertension’. Circulation Research 124 (6): 930–37. https://doi.org/10.1161/CIRCRESAHA.118.314487.

Tacutu, Robi, Daniel Thornton, Emily Johnson, Arie Budovsky, Diogo Barardo, Thomas Craig, Eugene Diana, et al. 2018. ‘Human Ageing Genomic Resources: New and Updated Databases’. Nucleic Acids Research 46 (D1): D1083–90. https://doi.org/10.1093/nar/gkx1042.

Tan, Nguan Soon, Liliane Michalik, Nicolas Di-Poï, Chuan Young Ng, Nicolas Mermod, Anita B. Roberts, Béatrice Desvergne, and Walter Wahli. 2004. ‘Essential Role of Smad3 in the Inhibition of Inflammation-Induced PPARbeta/Delta Expression’. The EMBO Journal 23 (21): 4211–21. https://doi.org/10.1038/sj.emboj.7600437.

Tatsumi, Yukako, and Takayoshi Ohkubo. 2017. ‘Hypertension with Diabetes Mellitus: Significance from an Epidemiological Perspective for Japanese’. Hypertension Research: Official Journal of the Japanese Society of Hypertension 40 (9): 795–806. https://doi.org/10.1038/hr.2017.67.

Tomomori, Shunsuke, Yukiko Nakano, Hidenori Ochi, Yuko Onohara, Akinori Sairaku, Takehito Tokuyama, Chikaaki Motoda, et al. 2018. ‘Maintenance of Low Inflammation Level by the ZFHX3 SNP Rs2106261 Minor Allele Contributes to Reduced Atrial Fibrillation Recurrence after Pulmonary Vein Isolation’. PloS One 13 (9): e0203281. https://doi.org/10.1371/journal.pone.0203281.

Touat-Hamici, Zahia, Henri Weidmann, Yuna Blum, Carole Proust, Hervé Durand, Francesca Iannacci, Veronica Codoni, et al. 2016. ‘Role of Lipid Phosphate Phosphatase 3 in Human Aortic Endothelial Cell Function’. Cardiovascular Research 112 (3): 702–13. https://doi.org/10.1093/cvr/cvw217.

Whitty, Christopher J. M., Carrie MacEwen, Andrew Goddard, Derek Alderson, Martin Marshall, Catherine Calderwood, Frank Atherton, et al. 2020. ‘Rising to the Challenge of Multimorbidity’. BMJ 368 (January): 16964. https://doi.org/10.1136/bmj.l6964.

Yang, Jian, Teresa Ferreira, Andrew P. Morris, Sarah E. Medland, Genetic Investigation of ANthropometric Traits (GIANT) Consortium, DIAbetes Genetics Replication and Meta-analysis (DIAGRAM) Consortium, Pamela A. F. Madden, et al. 2012. ‘Conditional and Joint Multiple-SNP Analysis of GWAS Summary Statistics Identifies Additional Variants Influencing Complex Traits’. Nature Genetics 44 (4): 369–75, S1-3. https://doi.org/10.1038/ng.2213.

Zaw, Khin Thet Thet, Noriko Sato, Shinobu Ikeda, Kaung Si Thu, Makiko Naka Mieno, Tomio Arai, Seijiro Mori, et al. 2017. ‘Association of ZFHX3 Gene Variation with Atrial Fibrillation, Cerebral Infarction, and Lung Thromboembolism: An Autopsy Study’. Journal of Cardiology 70 (2): 180–84. https://doi.org/10.1016/j.jjcc.2016.11.005.

