## Supplemental Information for "Integrative analysis of GWAS and co-localisation data suggests novel genes associated with age-related multimorbidity"

**Supplementary Information**

**SI Table 1)** For each age-related disease or trait, and for all traits combined (“Total”): the total number of GWAS studies, the number and proportion of these with summary statistics available; the minimum, median, and maximum number of individuals included in the GWAS studies; the total number of genome-wide significant trait-associated loci from these GWAS studies (“Total GWAS associations”); and the total number of independent genetic associations after clustering (“Total”), as well as the number of these clusters that included associations from multiple studies (“Replicated”) and the number that consisted of just one associated locus (“Unreplicated”).

|  | **N GWAS studies** | | **Study Population Sizes** | | |  | **N independent genetic associations (clusters)** | | |
| --- | --- | --- | --- | --- | --- | --- | --- | --- | --- |
| **Trait** | **Total** | **With summary statistics** | **Min** | **Max** | **Median** | **Total GWAS associations** | **Replicated** | **Unreplicated** | **Total** |
| adiponectin measurement | 9 | 0 (0%) | 3,310 | 67,739 | 29,347 | 48 | 9 | 19 | 28 |
| adult onset asthma | 7 | 1 (14%) | 327,253 | 585,812 | 327,253 | 336 | 30 | 234 | 264 |
| age-related macular degeneration | 16 | 4 (25%) | 2,721 | 399,047 | 33,976 | 108 | 13 | 50 | 63 |
| Alzheimer's disease | 62 | 11 (18%) | 2,222 | 1,462,121 | 61,571 | 265 | 34 | 85 | 119 |
| atherosclerosis | 15 | 8 (53%) | 31,211 | 397,126 | 218,792 | 142 | 24 | 64 | 88 |
| cartilage thickness measurement | 1 | 0 (0%) | 13,013 | 13,013 | 13,013 | 5 | 0 | 5 | 5 |
| chronic kidney disease | 24 | 4 (17%) | 4,863 | 625,219 | 67,093 | 92 | 16 | 54 | 70 |
| chronic obstructive pulmonary disease | 63 | 19 (30%) | 2,706 | 517,887 | 200,766 | 492 | 66 | 132 | 198 |
| Chronic pain | 1 | 0 (0%) | 387,649 | 387,649 | 387,649 | 39 | 0 | 39 | 39 |
| cognitive decline measurement | 2 | 0 (0%) | 3,946 | 3,946 | 3,946 | 16 | 0 | 16 | 16 |
| gait measurement | 2 | 0 (0%) | 2,946 | 450,967 | 450,967 | 72 | 0 | 72 | 72 |
| glaucoma | 37 | 8 (22%) | 4,365 | 402,223 | 218,792 | 518 | 79 | 98 | 177 |
| grip strength measurement | 12 | 2 (17%) | 4,608 | 359,729 | 359,704 | 547 | 138 | 208 | 346 |
| healthspan | 2 | 0 (0%) | 300,447 | 837,415 | 837,415 | 26 | 2 | 22 | 24 |
| heart disease | 146 | 59 (40%) | 2,845 | 1,030,836 | 296,525 | 2,449 | 315 | 333 | 648 |
| heel bone mineral density | 12 | 6 (50%) | 114,552 | 446,000 | 394,929 | 3,704 | 732 | 567 | 1,299 |
| hypertension | 29 | 11 (38%) | 4,965 | 517,472 | 361,141 | 1,019 | 181 | 229 | 410 |
| idiopathic pulmonary fibrosis | 1 | 0 (0%) | 11,259 | 11,259 | 11,259 | 13 | 0 | 13 | 13 |
| lean body mass | 24 | 12 (50%) | 2,207 | 354,808 | 354,736 | 7,421 | 1,045 | 223 | 1,268 |
| Lewy body dementia | 3 | 0 (0%) | 3,526 | 3,526 | 3,526 | 3 | 1 | 0 | 1 |
| lipid measurement | 690 | 10 (1%) | 2,011 | 859,386 | 127,326 | 7,210 | 682 | 1,292 | 1,974 |
| longevity | 25 | 8 (32%) | 4,477 | 837,415 | 482,000 | 160 | 21 | 38 | 59 |
| metabolic syndrome | 14 | 0 (0%) | 2,554 | 291,107 | 291,107 | 126 | 13 | 84 | 97 |
| obesity | 21 | 8 (38%) | 2,928 | 408,961 | 204,498 | 137 | 26 | 29 | 55 |
| osteoarthritis | 44 | 27 (61%) | 6,523 | 613,790 | 393,873 | 302 | 45 | 64 | 109 |
| osteoporosis | 8 | 4 (50%) | 3,569 | 407,763 | 361,141 | 20 | 3 | 12 | 15 |
| Parkinson's disease | 21 | 2 (10%) | 2,755 | 482,730 | 417,508 | 244 | 47 | 83 | 130 |
| rheumatoid arthritis | 54 | 11 (20%) | 5,683 | 369,964 | 80,799 | 776 | 126 | 231 | 357 |
| stroke | 44 | 11 (25%) | 4,348 | 922,140 | 100,321 | 168 | 37 | 55 | 92 |
| type II diabetes mellitus | 90 | 16 (18%) | 4,347 | 1,407,282 | 659,316 | 3,062 | 484 | 541 | 1,025 |
| Total | 1394 | 231 (17%) | 2,011 | 1,462,121 | 35,634 | 28,797 |  |  |  |

**SI Table 2**

CSV file containing EFO codes and descriptions for the age-related diseases and traits, including descendant terms, the corresponding number of GWAS studies and genome-wide significant associations from the studies, and the proportion of studies for which full summary statistics are available. This table includes diseases and traits that were included in the curated list but for which there were no GWAS studies available with at least 2000 individuals.

**SI Table 3**

GeneOntology term mappings for each of the Hallmarks of Ageing, the number of descendant terms in the ontology, and the number of TargetAge genes annotated with each term (including descendent terms) and each Hallmark overall.

| **Hallmark** | **GO Term ID** | **Go Term Name** | **N descendant terms** | **N TargetAge Genes** | **N TargetAge Genes** |
| --- | --- | --- | --- | --- | --- |
| **Cellular senescence** | GO:0090398 | cellular senescence | 7 | 17 | 17 |
| **Proteostasis** | GO:0006457 | protein folding | 23 | 5 | 70 |
|  | GO:0050821 | protein stabilization | 1 | 9 |  |
|  | GO:0030163 | protein catabolic process | 145 | 64 |  |
|  | GO:0010498 | proteasomal degradation | 51 | 25 |  |
|  | GO:0061684 | chaperone-mediated autophagy | 7 | 2 |  |
|  | GO:0061077 | chaperone-mediated protein folding | 8 | 2 |  |
| **Telomere maintenance** | GO:0000723 | telomere maintenance | 54 | 7 | 7 |
| **Nutrient signalling** | GO:0031667 | response to nutrient levels | 190 | 69 | 72 |
|  | GO:0009594 | detection of nutrient | 2 | 0 |  |
|  | GO:0048009 | IGF receptor signaling pathway | 4 | 5 |  |
| **Mitochondrial genome maintenance** | GO:0000002 | mitochondrial genome maintenance | 10 | 0 | 0 |
| **Epigenetic alteration** | GO:0044728 | DNA methylation or demethylation | 29 | 8 | 58 |
|  | GO:0016570 | histone modification | 215 | 45 |  |
|  | GO:0006338 | chromatin remodeling | 54 | 24 |  |
| **Inflammation** | GO:0006954 | inflammatory response | 174 | 65 | 65 |
| **Immune response** | GO:0006955 | immune response | 661 | 92 | 92 |

**SI Table 4**

TargetAge targets with high-quality chemical probes available and no clinical precedence, the name, source, and URL of the probe.

| **Target ID** | **Target Symbol** | **Probe ID** | **Source** | **URL** |
| --- | --- | --- | --- | --- |
| ENSG00000119778 | ATAD2B | BAY-850 | Bromodomains chemical toolbox | https://www.nature.com/articles/s41467-019-09672-2#article-info |
|  |  |  | Chemical Probes.org (legacy) | https://new.chemicalprobes.org/?q=BAY-850 |
|  |  |  | SGC Probes | http://www.thesgc.org/chemical-probes/BAY-850 |
|  |  | GSK8814 | Bromodomains chemical toolbox | https://www.nature.com/articles/s41467-019-09672-2#article-info |
|  |  |  | SGC Probes | http://www.thesgc.org/chemical-probes/GSK8814 |
|  |  |  | Chemical Probes.org (legacy) | https://new.chemicalprobes.org/?q=GSK8814 |
| ENSG00000171634 | BPTF | NVS-BPTF-1 | SGC Probes | http://www.thesgc.org/chemical-probes/NVS-BPTF-1 |
|  |  | TP-238 | SGC Probes | http://www.thesgc.org/chemical-probes/TP-238 |
| ENSG00000104885 | DOT1L | canSAR828356 | Chemical Probes.org (legacy) | https://new.chemicalprobes.org/?q=canSAR828356 |
|  |  |  | Protein methyltransferases chemical toolbox | https://www.nature.com/articles/s41467-018-07905-4 |
|  |  | canSAR851884 | Chemical Probes.org (legacy) | https://new.chemicalprobes.org/?q=canSAR851884 |
|  |  |  | Probe Miner | http://probeminer.icr.ac.uk/#/Q8TEK3 |
|  |  | canSAR856426 | Chemical Probes.org (legacy) | https://new.chemicalprobes.org/?q=canSAR856426 |
|  |  |  | Probe Miner | http://probeminer.icr.ac.uk/#/Q8TEK3 |
| ENSG00000198945 | L3MBTL3 | UNC1215 | Nature Chemical Biology Probes | http://www.nature.com/nchembio/chemical_probes.html |
|  |  |  | SGC Probes | http://www.thesgc.org/chemical-probes/UNC1215 |
|  |  |  | Chemical Probes.org (legacy) | https://new.chemicalprobes.org/?q=UNC1215 |
| ENSG00000168906 | MAT2A | AG-270 | Chemical Probes.org (legacy) | https://new.chemicalprobes.org/?q=AG-270 |
| ENSG00000078142 | PIK3C3 | SAR405 | Nature Chemical Biology Probes | http://www.nature.com/nchembio/chemical_probes.html |
| ENSG00000198890 | PRMT6 | SGC6870 | SGC Probes | http://www.thesgc.org/chemical-probes/SGC6870 |
|  |  | canSAR1301406 | Protein methyltransferases chemical toolbox | https://www.nature.com/articles/s41467-018-07905-4 |
|  |  |  | Chemical Probes.org (legacy) | https://new.chemicalprobes.org/?q=canSAR1301406 |
| ENSG00000142178 | SIK1 | MRIA9 | SGC Probes | http://www.thesgc.org/chemical-probes/MRIA9 |
|  |  | canSAR2945471 | Chemical Probes.org (legacy) | https://new.chemicalprobes.org/?q=canSAR2945471 |
| ENSG00000178950 | GAK | CA93.0 | SGC Probes | http://www.thesgc.org/chemical-probes/CA93.0 |
| ENSG00000137764 | MAP2K5 | canSAR326840 | Chemical Probes.org (legacy) | https://new.chemicalprobes.org/?q=canSAR326840 |
| ENSG00000160584 | SIK3 | MRIA9 | SGC Probes | http://www.thesgc.org/chemical-probes/MRIA9 |
|  |  | canSAR2945471 | Chemical Probes.org (legacy) | https://new.chemicalprobes.org/?q=canSAR2945471 |

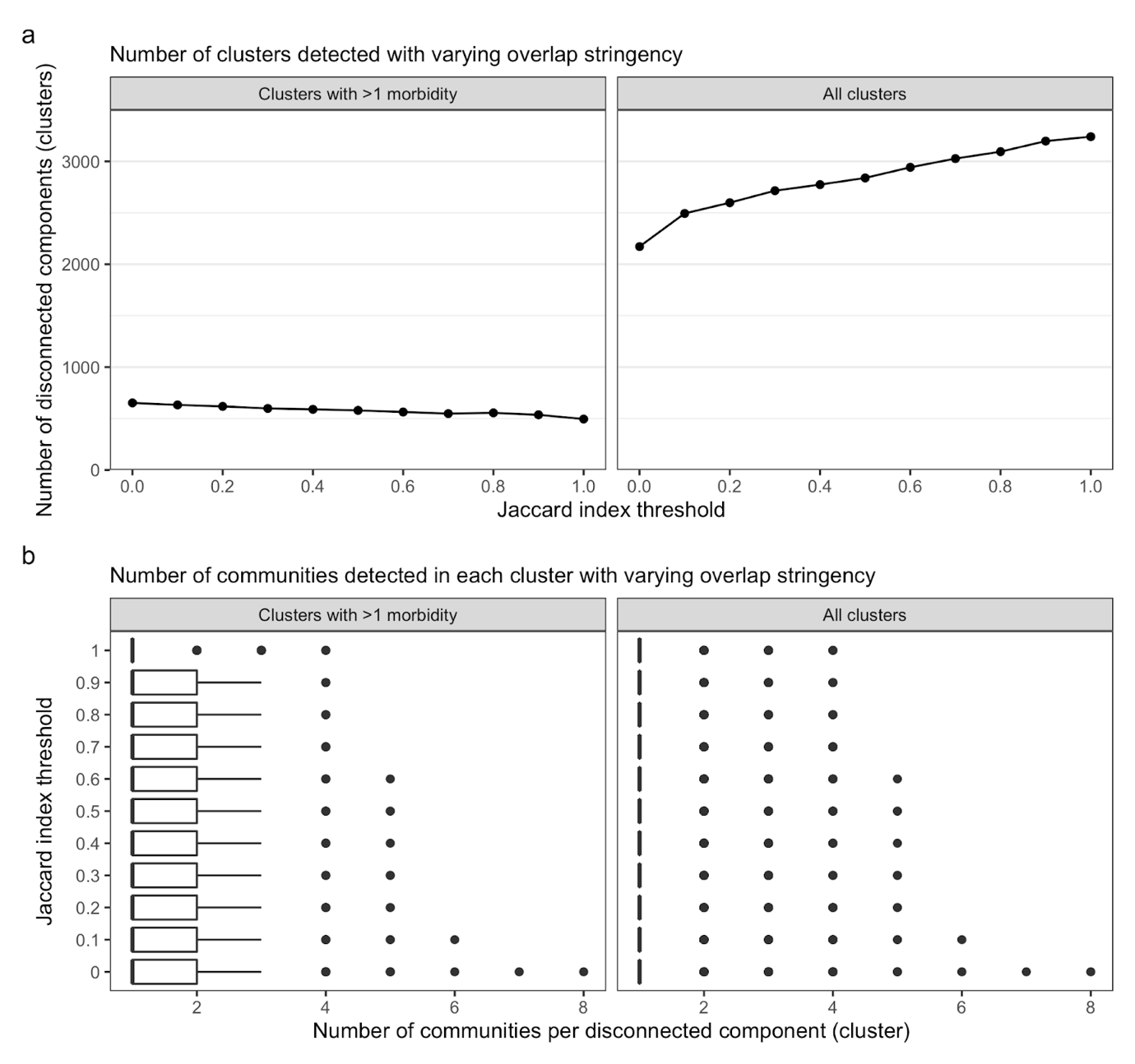

**SI Figure 1) The effect of using a more stringent overlap criteria on the number of clusters and sub-cluster communities in the graph**. The minimum overlap between the tag variants of two genetic associations, defined using the Jaccard Index (i.e. the ratio of the intersection over the union), required to define an edge between two genetic associations was varied from >0 to 1, to compare the number of a) clusters and b) sub-cluster communities identified in the resulting graphs.

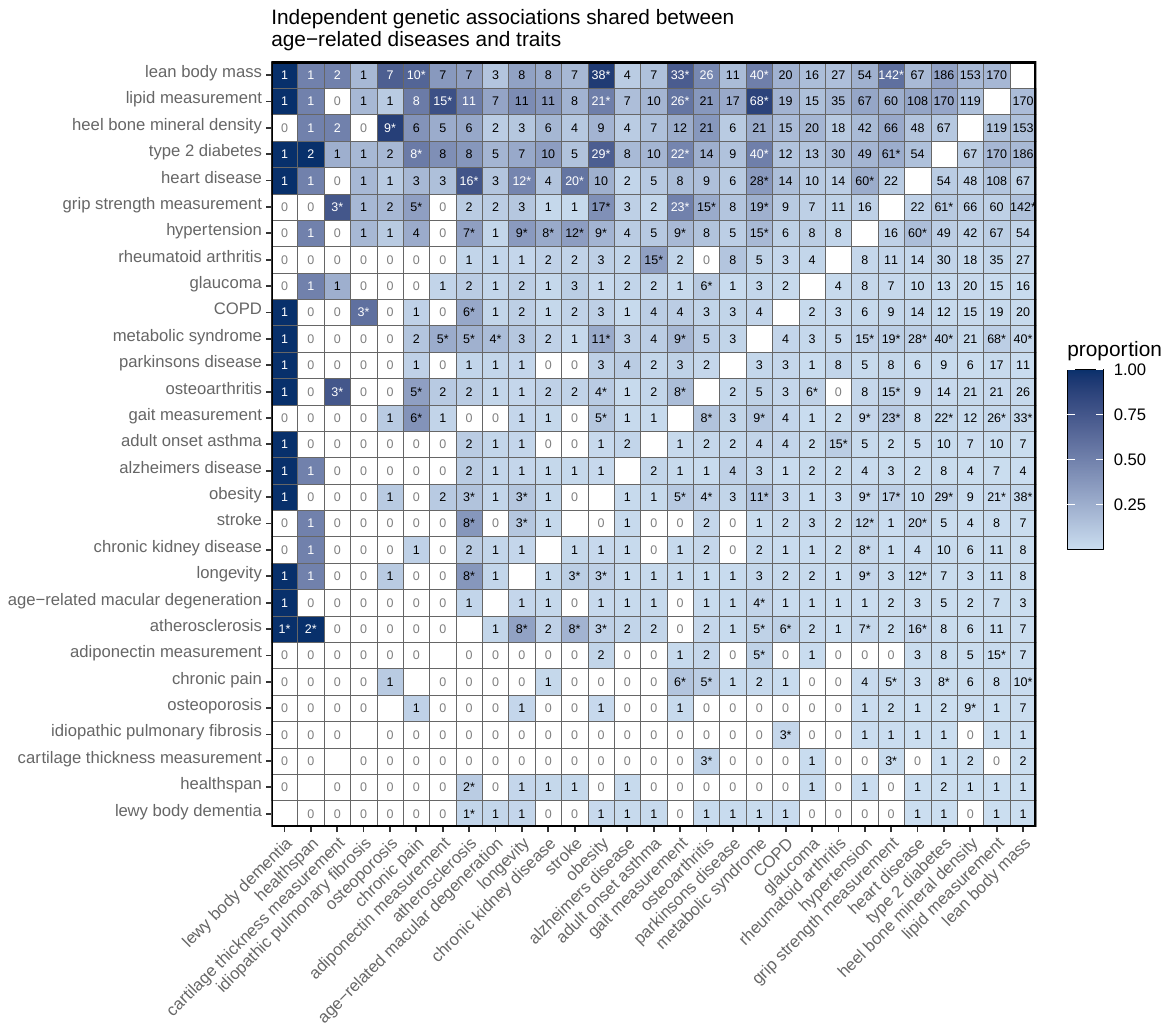

**SI Figure 2) The number and proportion of independent genetic associations shared between age-related diseases and traits.** Proportions are relative to the total number of independent genetic associations identified for the trait on the horizontal axis; darker blue indicates a greater proportion of shared genetic associations. Cells marked with an asterisk are statistically significant after adjusting for multiple testing of all pairwise combinations of traits. Traits are ordered by the total number of implicated genetic association clusters.
